# A diagnostic model based on differential whole-brain dynamics for distinguishing neuropsychiatric symptom and cognitive impairment

**DOI:** 10.64898/2026.04.27.26351804

**Authors:** Lihong Huang, Mengyu Yan, Zhangjing Deng, Yang Lü, Weihua Yu

**Affiliations:** Institutes of Neuroscience, Chongqing Medical University, Chongqing 400016, China; Department of Geriatrics, The First Affiliated Hospital of Chongqing Medical University, Chongqing 400016, China

**Keywords:** Alzheimer’s disease, cognitive impairment, neuropsychiatric symptoms, EEG microstates, LASSO

## Abstract

**Objectives:** Neuropsychiatric symptoms (NPS) are prevalent in individuals of cognitive impairement (CI). However, the similarities and disparatenesses in whole-brain dynamics between individuals of CI and NPS are controversy. Electroencephalography (EEG) microstates reflect the whole-brain dynamics. This study aimed to investigate the differential EEG microstates parameters between CI and NPS and to construct related diagnostic model.

**Methods/design:** This study was a cross-sectional study. Clinical and EEG data were collected, and an EEG microstate analysis were performed. The Least absolute shrinkage and selection operation (LASSO) regression model was used to identify significant differential EEG microstates parameters between CI and NPS and to construct a diagnostic model. The model performance was tested by the receiver operating characteristic curve (ROC).

**Results:** This study enrolled 78 participants. A total of 36 EEG microstates parameters were identified and included in the differential analysis. In the LASSO regression model, 4 significant differential EEG microstates parameters were selected, including the duration of class C, TPAB, TP_BA_, and TP_DC_. The ROC results showed that the diagnostic model for distinguishing NPS patients from CI patients achieved an area under the curve (AUC) of 0.905(95% CI: 0.784-1.000), with a sensitivity of 100.0% and a specificity of 76.9%.

**Conclusions:** The diagnostic model based on EEG microstate parameters showed a good performance for differentiating NPS patients from CI patients.

## 1. Background

Dementia is prevalent in older adults[1]. Cognitive impairment (CI) is the early salient clinical feature of dementia [2]. Additionally, patients additionally exhibit behavioral or neuropsychiatric symptoms (NPS) changes. NPS refers to behavioral, affective, and personality changes that can be attributed to underlying neurodegenerative disease [3]. Recent studies have indicated that NPS are prevalent not only in individuals in the early stages of AD, but also in many instances preceding the onset of identifiable neurocognitive changes [4-6]. An increase of NPS in the early stages of AD is associated with more rapid cognitive decline, increased caregiver burden, institutionalization, and mortality [5, 7]. Therefore, NPS are considered diagnostic and prognostic indicators of AD-associated dementia. Accurate approaches for assessing NPS are important for accurate differential diagnosis, disease management, and understanding the neurobiological underpinnings of behavioral changes in patients [8].

Currently, there are several tools available for assessing the presence and severity of CI and of NPS in AD patients, respectively. Among them, the most widely used is the clinical scale, which is an informant-based questionnaire [9]. However, this method is subjective and limited to identifying clinical phenotypes only. Thus, this approach cannot help us fully understand the underlying brain dynamics mechanism. Some studies have suggested that there were shared underlying mechanism between CI and NPS [10], while other studies also indicated that differing functional connectivity abnormalities in CI and NPS[11]. Thus, there are controversy of similarities and disparatenesses in brain dynamics between individuals of CI and NPS, which need to be further explored.

Electroencephalography (EEG) is a useful tool for evaluating brain function in AD patients [12]. Previous studies have shown that EEG characteristics play an important role in the clinical diagnosis of AD [13, 14]. Especially, resting-state EEG data has its advantages in clinical settings, such as being noninvasive, reproducible, cost effective and minimal requirements for patient participation [15]. Furthermore, compared with other EEG quantification parameters, such as power spectral density, event-related potentials and nonlinear dynamic parameters, EEG microstate parameters can provide insight into the the whole-brain dynamics changes across different timescales. Studying EEG microstate dynamics on a sub-second timescale can therefore provide information about brain dynamics with implications for dynamic cognitive processes [16]. Therefore, among the various EEG quantification parameters, EEG microstates can be useful for assessing NPS of AD patients. EEG microstates parameters has explored in many other neurological and psychiatric disorders, including schizophrenia[17], bipolar disorder and depression[18], and epilepsy[19], but their roles in CI and NPS phenotype of AD patients have not yet been explored.

This study aims to investigate the differential EEG microstates parameters among NPS patients, CI patients and healthy control (HC) subjects. It also aimes to apply the Least absolute shrinkage and selection operation (LASSO) - logistic regression to establish a diagnostic model based on EEG microstate for distinguishing NPS and CI, which could help provide an effective reference for the identification of NPS and for the exploration of its potential underlying mechanisms of whole-brain dynamics alteration.

## 2. Methods

### 2.1 Study Design

This is a cross-sectional study. Patients were recruited from the Memory Clinic, Department of Geriatrics, The First Affiliated Hospital of Chongqing Medical University between January 2022 and June 2023. The HC subjects were recruited from the community. The HC subjects were recruited through public advertisements at memory clinics, local associations for elderly people, and an online recruitment site for trial subjects. The study protocol was approved by the Medical Ethics Committee of the First Affiliated Hospital of the Chongqing Medical University.

### 2.2 Participants

The inclusion criteria were as follows: (1) age ≥ 60 years and (2) met the clinical diagnostic criteria for CI and NPS. The diagnostic criteria for CI met the Winblad consensus criteria [20] and NIA/AA diagnostic guidelines, respectively [21]. NPS was diagnosed based on the NPI. The inclusion criteria for HCs were as follows: (1) age ≥ 60 years, (2) Chinese version of the Mini-Mental State Examination (CMMSE) score ≥ 27, (3) Clinical Dementia Rating Scale (CDR) score = 0, (4) normal performance on standardized neuropsychological tests and had or lacked cognitive complaints or concerns during the structured interview, and (5) no neurological disorders that affected cognitive function.

The exclusion criteria were as follows: (1) a history of psychiatric illness, severe head trauma or brain injury, multiple sclerosis, or a history of alcohol abuse or drug abuse within the past five years; (2) intellectual disabilities leading to cognitive decline; (3) CI caused by other neurological diseases, such as trauma, stroke, tumors, Parkinson’s disease, encephalitis, epilepsy, or other types of dementia, such as frontotemporal dementia, Lewy Body Dementia, and vascular dementia; (4) CI caused by severe anemia, thyroid diseases, and other systemic illnesses; (5) skull defects, malignant tumors, or other serious illnesses; and (6) inability to complete neuropsychological tests or incomplete clinical data.

### 2.3 Patient group

Patients were divided into three groups, including NPS, CI and HC groups, according to their clinical CMMSE, CDR and NPI scores. The CI group included patients with mild cognitive impairment and mild dementia. Due to all NPS patients exhibit CI in this study, therefore, the CI patients were further divided into a CI without NPS (CI) group and CI combined with NPS (NPS) group according to their NPI scores.

### 2.4 Data collection

#### 2.4.1 Clinical data collection

Clinical data collection, including demographic characteristics, medical history, drug use, brain magnetic resonance imaging (MRI) (including medial temporal atrophy (MTA) and Fazekas), and neuropsychological test results, was conducted by a professional researcher. The drugs used included anti-dementia and antipsychotic drugs. The antidementia drugs used included cholinesterase inhibitors and N-methyl-d-aspartate (NMDA) receptor antagonists. The antipsychotic drugs reported by the participants included anticonvulsants, antidepressants, antihistamines, antipsychotics, anxiolytics, benzodiazepines, cholinesterase inhibitors, hypnotics, mood stabilizers, NMDA-receptor antagonists, and stimulants [22]. All participants underwent thorough neuropsychological tests, including the CMMSE [23] and CDR (score range: 0–3) [24], to assess global cognitive ability and NPI for NPS. The CMMSE scores ranged from 0 (severe impairment) to 30 (no impairment). The CDR ranges from 0.0 (normal), 0.5 (mild cognitive impairment), 1.0 (mild dementia), 2.0 (moderate dementia), to 3.0 (severe dementia). The NPI was used to assess NPS [9]. The NPI relied on a caregiver/informant report of the presence, severity, and distress caused by 12 NPS that were evident within the past month. The symptoms assessed included delusions, hallucinations, agitation/aggression, depression/dysphoria, anxiety, elation/euphoria, apathy/indifference, disinhibition, irritability/lability, motor disturbances, nighttime behaviors, and appetite/eating problems. The composite score for each NPI subscale ranged from 0 (no NPS) to 12, with the total composite score ranging from 0 (no NPS) to 144. The patients underwent neuropsychological assessment via face-to-face interviews. The assessments were performed by a trained nurse or an experienced doctor.

#### 2.4.2 EEG data

In this study, the EEG signals were all scalp electroencephalographic signals recorded by an EEG monitoring system (N1M1-neuracle). Standard for EEG recording: Before EEG recording, electrodes were installed according to the international 10/20 system, which included 19 recording channels (FP1/FP2, F3/F4, C3/C4, P3/P4, O1/O2, F7/F8, T3/T4, T5/T6, Fz, Cz, Pz), 2 earlobe reference electrodes (A1 and A2), 1 common reference electrode (reference), and 1 ground electrode (GND). During the installation process, a conductive gel was used to maintain the impedance at less than 5 kΩ. The participants were instructed to wash their hair the day before formal collection of EEG data to ensure good sleep. Before the collection of EEG data, the participants were asked to rest quietly for 5 min. The EEG signals were collected in the resting state, meaning that the patients rested quietly for a total duration of 10 min. The EEG data were collected using the following settings: Montage, standard 10–20 electrode placement; sampling rate, 512 Hz; notch filter, 49–51 Hz; and bandpass filter: 0.3 70 Hz. Sensitivity: 7 μV/mm; EEG paper speed: 30 mm/s [25].

#### 2.4.3 Processing and Quantitative Analysis of EEG Data

##### (1) EEG Data Preprocessing

EEG data were imported into MATLAB (MathWorks, v2013b) using the EEGLAB toolbox [26]. The EEG length of each patient was 10 minutes, and only segments with closed eyes were selected for analysis. (1) The electrodes were computationally located on the scalp using a standard 10–20 electrode model. (2) Excessive channels were removed and the data were band-pass filtered from 0.5-40 Hz. The data were filtered at 45–55 Hz. (3) Subsequently, the filtered data were resampled at 250 Hz and segmented for 2 s. (4) The processed data were rereferenced using earlobe electrodes (A1/A2). (5) EEGs were visually inspected and epochs with excessive noise or artifacts (chewing, swallowing, and muscle artifacts) were removed. Channels with excessive noise, drift, or poor connections were interpolated using spherical interpolation. Patients with ≥ three interpolated electrodes were excluded from analysis. (6) The EEGs were visually inspected again and epochs with excessive noise or artifacts were removed. The investigator who performed preprocessing was blinded to the diagnosis.

##### (2) Analysis of EEG microstate parameters

Before performing the EEG microstate analysis, the data were filtered at 2-20 Hz using the same settings as mentioned above. The EEG microstate analysis was performed using the Microstate EEGLAB Toolbox. First, global field power peaks were extracted. All GFP peaks from all the subjects were aggregated into one file before segmentation to maximize the similarity between the microstates to which they would be assigned. For segmentation, a modified atomized and agglomerated hierarchical clustering algorithm was used, which was specifically designed for EEG clustering [27]. Four classes of microstates (Classes A, B, C, and D) were previously reported as the most common and reproducible [28]. The number of repetitions was set to 50 and the maximum number of iterations was set to 1000. The global maps (**see Figure 1**) were then back-fitted to each EEG file by labeling each EEG segment with the most familiar class of microstates. After back-fitting the global maps, the duration, occurrence, coverage, and transitional properties (TP) of the EEG files were calculated. In this study, △TP is defined as the expected and calculated TP.

**Figure 1.**
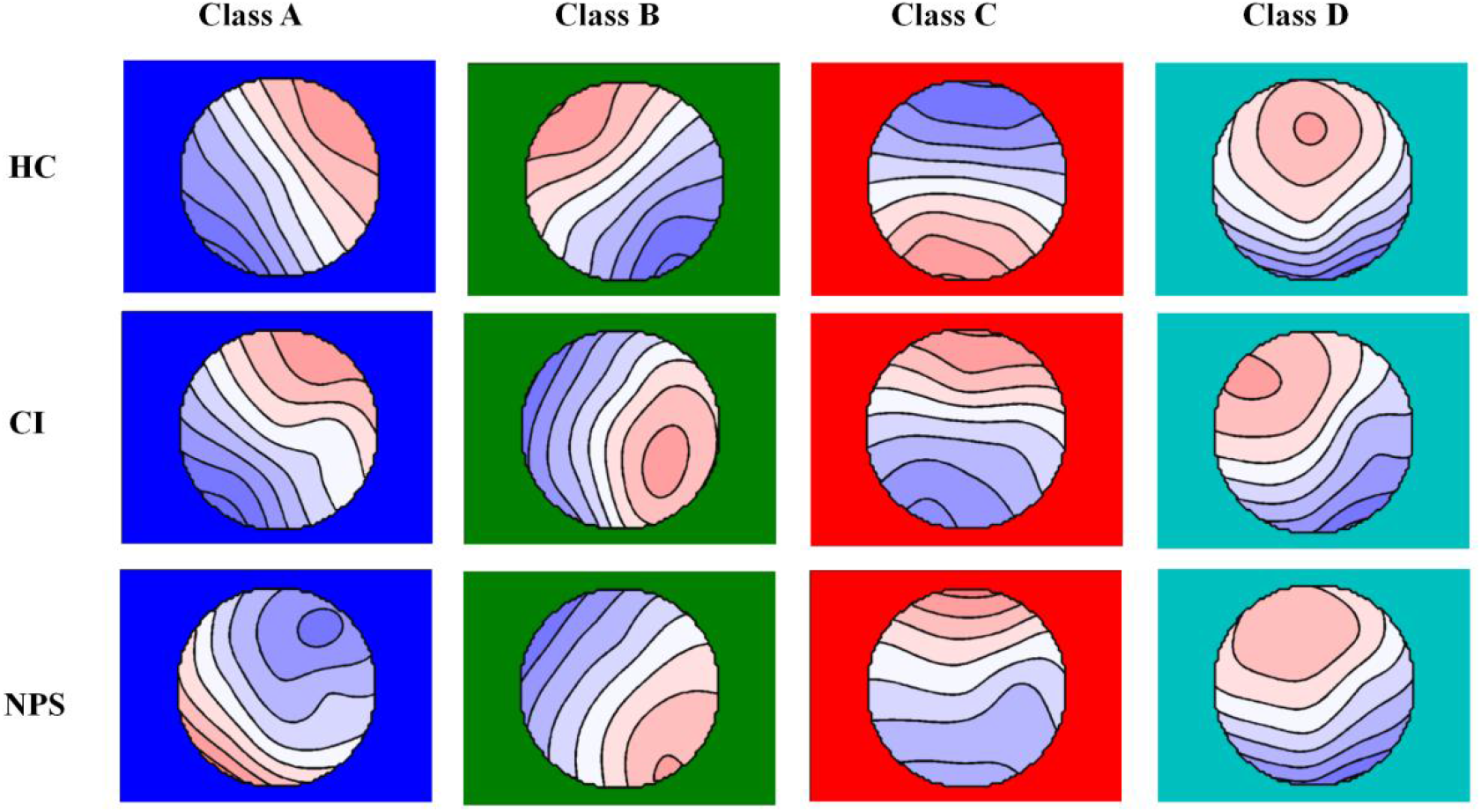
EEG microstate topolograph of 3 groups.

### 2.5 Statistical analysis

All data analyses were performed using SPSS (version 23.0) and R (version 4.05). Quantitative data are presented as the mean ± standard deviation (SD), and qualitative data are presented as frequencies and percentages (n, %). Analysis of Variance (ANOVA) was used for mean comparison of normal data, whereas the Kruskal-Wallis H test was used for non-normal data distribution comparison. A chi-square test was conducted for the qualitative data. Based on the glmnet package, the least absolute shrinkage and selection operator (LASSO) was used to further select EEG variables that showed differences in univariate analysis. Finally, models were constructed to diagnose CI patients and NPS patients using EEG variables selected by LASSO regression. These models were evaluated using receiver operating characteristic (ROC) curves and sensitivity and specificity metrics. Statistical significance was set at P < 0.05 (two-tailed) was considered as statistical significance.

## 3. Results

### 3.1 General Characteristics of Participants

This study included 78 patients who met the following inclusion criteria: 21 NPS patients, 37 CI patients, and 20 HC subjects. **Table 1** shows a comparison of patient baseline characteristics, including demographic characteristics, clinical data, CMMSE scores, and CDR scores, among the three groups. There were no statistically significant differences among three groups in terms of age, sex, education, medical history, or drug use (P>0.05). The clinical data of CMMSE score, CDRs, MTA and Fazekas showed no statistically significant differences between CI and NPS groups (P>0.05).

**Table 1.** Comparison of baseline characteristics among 3 groups.

| Variables |  | HC group<br>(N=20) | CI group<br>(N=37) | NPS group<br>(N=21) | P values |
| --- | --- | --- | --- | --- | --- |
| Age |  | 69.11±7.12 | 71.77±5.96 | 72.15± 5.60 | 0.38 |
| Gender N(%) | Male | 9 (0.45) | 22 (0.38) | 11 (0.29) | 0.55 |
|  | Female | 11 (0.55) | 36 (0.62) | 28 (0.71) |  |
| Education |  | 9.78± 4.41 | 10.54± 3.95 | 8.62± 3.43 | 0.41 |
| Medical History N(%) | No | 19 | 31 | 18 | 0.68 |
|  | Yes | 1 | 6 | 3 |  |
| Use of drugs N(%) | No | 19 | 33 | 20 | 0.62 |
|  | Yes | 1 | 4 | 1 |  |
| CMMSE |  | 27.61± 1.72 | 20.11±5.26 | 19.85± 8.59 | 0.29 |
| CDR |  | 0 | 0.79± 0.30 | 0.86± 0.39 | 0.40 |
| MTA |  | -- | 2.12± 0.67 | 2.1± 0.50 | 0.95 |
| Fezakas |  | -- | 1.36± 0.32 | 1.32± 1.00 | 0.82 |

### 3.2 The Results of EEG Microstate Characteristics

**Table 2** shows a comparison of the EEG microstate parameters among the three groups. There were 8 diferential EEG microstate parameters between CI and NPS groups (P < 0.05). Among these, compared to those in CI group, duration of class C (0.06±0.01 vs. 0.07±0.01, P=0.03), TP_BA_ (0.11±0.02 vs. 0.12±0.05, P <0.01), TP_DC_ (0.06 ± 0.02 vs.0.08±0.03, P <0.01), △TP_CD_ (0.007 ± 0.001 vs. 0.009 ± 0.002, P <0.01), and △TP_DC_ (0.007 ± 0.001 vs. 0.008 ± 0.002, P <0.01) were significantly higher in the NPS group. However, compared to those in the CI group, the TP_AB_ (0.08±0.02 vs. 0.12±0.04), △TP_AB_ (0.08±0.01 vs. 0.10 ± 0.02) and △TP_BA_ (0.008 ± 0.001 vs. 0.010± 0.002) were decreased in the NPS group.

**Table 2.** Comparison of EEG microstate parameters of 3 groups.

| Variables | HC group<br>(N=20) | CI group<br>(N=37) | NPS group<br>(N=21) | P values | Post hoc |
| --- | --- | --- | --- | --- | --- |
| Duration_A | 0.07± 0.01 | 0.07± 0.01 | 0.07± 0.01 | 0.61 |  |
| Duration_B | 0.07± 0.01 | 0.07± 0.01 | 0.07± 0.01 | 0.30 |  |
| Duration_C | 0.06± 0.02 | 0.06± 0.01 | 0.07± 0.01 | 0.041<sup>c</sup> | NPS>CI |
| Duration_D | 0.06± 0.02 | 0.06± 0.01 | 0.07± 0.01 | 0.07 |  |
| Occurrence_A | 4.37± 1.20 | 3.84± 0.76 | 3.51± 0.83 | 0.06 |  |
| Occurrence_B | 4.29± 0.97 | 4.12± 0.59 | 3.62± 0.85 | 0.07 |  |
| Occurrence_C | 3.26± 0.65 | 3.41± 0.64 | 3.74± 0.74 | 0.19 |  |
| Occurrence_D | 3.56± 0.64 | 3.25± 0.72 | 3.58± 0.41 | 0.40 |  |
| Contribution_A | 0.30± 0.10 | 0.27± 0.06 | 0.24± 0.10 | 0.20 |  |
| Contribution_B | 0.28± 0.07 | 0.29± 0.05 | 0.25± 0.10 | 0.13 |  |
| Contribution_C | 0.20± 0.07 | 0.23± 0.06 | 0.26± 0.08 | 0.06 |  |
| Contribution_D | 0.22± 0.09 | 0.21± 0.06 | 0.25± 0.05 | 0.24 |  |
| TP A→B | 0.12± 0.04 | 0.11± 0.02 | 0.08± 0.02 | <0.01<sup>bc</sup> | HC>NPS,<br>CI>NPS |
| TP A→C | 0.08± 0.02 | 0.08± 0.02 | 0.08± 0.03 | 0.44 |  |
| TP A→D | 0.08± 0.03 | 0.07± 0.02 | 0.08± 0.03 | 0.52 |  |
| TP B→A | 0.12± 0.05 | 0.11± 0.02 | 0.08± 0.02 | <0.01<sup>bc</sup> | CI>NPS,<br>HC>NPS |
| TP B→C | 0.07± 0.03 | 0.09± 0.03 | 0.09± 0.03 | 0.15 |  |
| TP B→D | 0.08± 0.02 | 0.08± 0.02 | 0.08± 0.04 | 0.73 |  |
| TP C→A | 0.07± 0.02 | 0.08± 0.02 | 0.09± 0.03 | 0.37 |  |
| TP C→B | 0.07± 0.02 | 0.08± 0.02 | 0.08± 0.02 | 0.13 |  |
| TP C→D | 0.07± 0.04 | 0.07± 0.02 | 0.09± 0.03 | 0.09 |  |
| TP D→A | 0.08± 0.03 | 0.07± 0.02 | 0.08± 0.03 | 0.63 |  |
| TP D→B | 0.08± 0.02 | 0.09± 0.02 | 0.08± 0.04 | 0.54 |  |
| TP D→C | 0.07± 0.04 | 0.06± 0.02 | 0.08± 0.03 | <0.046<sup>c</sup> | NPS>CI |
| △TPA→B | 0.11± 0.04 | 0.10± 0.02 | 0.08± 0.01 | <0.01<sup>bc</sup> | CI>NPS,<br>HC>NPS |
| △TPA→C | 0.008± 0.002 | 0.008± 0.002 | 0.008± 0.002 | 0.73 |  |
| △TPA→C | 0.009± 0.003 | 0.008± 0.002 | 0.008± 0.003 | 0.45 |  |
| △TPB→A | 0.011± 0.003 | 0.010± 0.002 | 0.008± 0.001 | <0.01<sup>bc</sup> | CI>NPS,<br>HC>NPS |
| △TPB→C | 0.008± 0.002 | 0.009± 0.002 | 0.009± 0.002 | 0.41 |  |
| △TPB→D | 0.009± 0.001 | 0.009 ±0.002 | 0.009± 0.003 | 0.37 |  |
| △TPC→A | 0.007± 0.001 | 0.008 ±0.002 | 0.008± 0.002 | 0.10 |  |
| △TPC→B | 0.007± 0.002 | 0.009 ±0.002 | 0.009± 0.002 | 0.26 |  |
| △TPC→D | 0.007± 0.003 | 0.007 ±0.001 | 0.009± 0.002 | <0.01<sup>bc</sup> | NPS>CI,<br>NPS>HC |
| △TPD→A | 0.008± 0.003 | 0.007 ±0.002 | 0.008± 0.002 | 0.63 |  |
| △TPD→B | 0.008± 0.002 | 0.008 ±0.002 | 0.008± 0.003 | 0.91 |  |
| △TPD→C | 0.007± 0.003 | 0.007 ±0.001 | 0.008± 0.002 | <0.01<sup>bc</sup> | NPS>HC,<br>NPS>CI |

a.comaparion between HC and CI group, P < 0.05 after adjustment. b.comaparion between HC and NPS group, P<0.05 after adjustment. c. comaparion between CI and NPS group, P<0.05 after adjustment. TP= transitional probabilities.

Furthermore, there were 6 differential EEG microstate parameters between NPS and HC groups (P < 0.05). Among these, compared to those in HC group, the TP_BA_ (0.12±0.05 vs. 0.08±0.02, P <0.01), △TP_CD_(0.009± 0.002 vs. 0.007±0.003, P <0.01), △TP_DC_(0.008± 0.002 vs. 0.007±0.003, P <0.01) were significantly higher in the NPS group. However, compared to those in the HC group, TP_AB_(0.08 ± 0.02 vs. 0.12 ± 0.04, P <0.01), △TP_AB_(0.08± 0.01VS. 0.11± 0.04, P <0.01), △TP_BA_(0.008± 0.001 VS. 0.011± 0.003, P <0.01) were decreased in the NPS group. There were no significant differences in the remaining parameters among the 3 groups (all p >0.05).

### 3.3 Construction of diagnostic model for distinguishing NPS and CI

#### 3.3.1 Identification of a diagnostic model based on LASSO regression

Based on the previous analysis, 8 differential EEG microstates (P<0.05) were selected in LASSO regression. When the LASSO regression model achieved optimal predictive performance, with lambda (min) equal to 0.0157 (logλ = -4.15) (**Figure 2A-2B**), 4 EEG microstate parameters were identified, including the duration of class C, TP_DC_, TP_AB_ and TP_BA_ (**Figure 3**).

**Figure 2.**
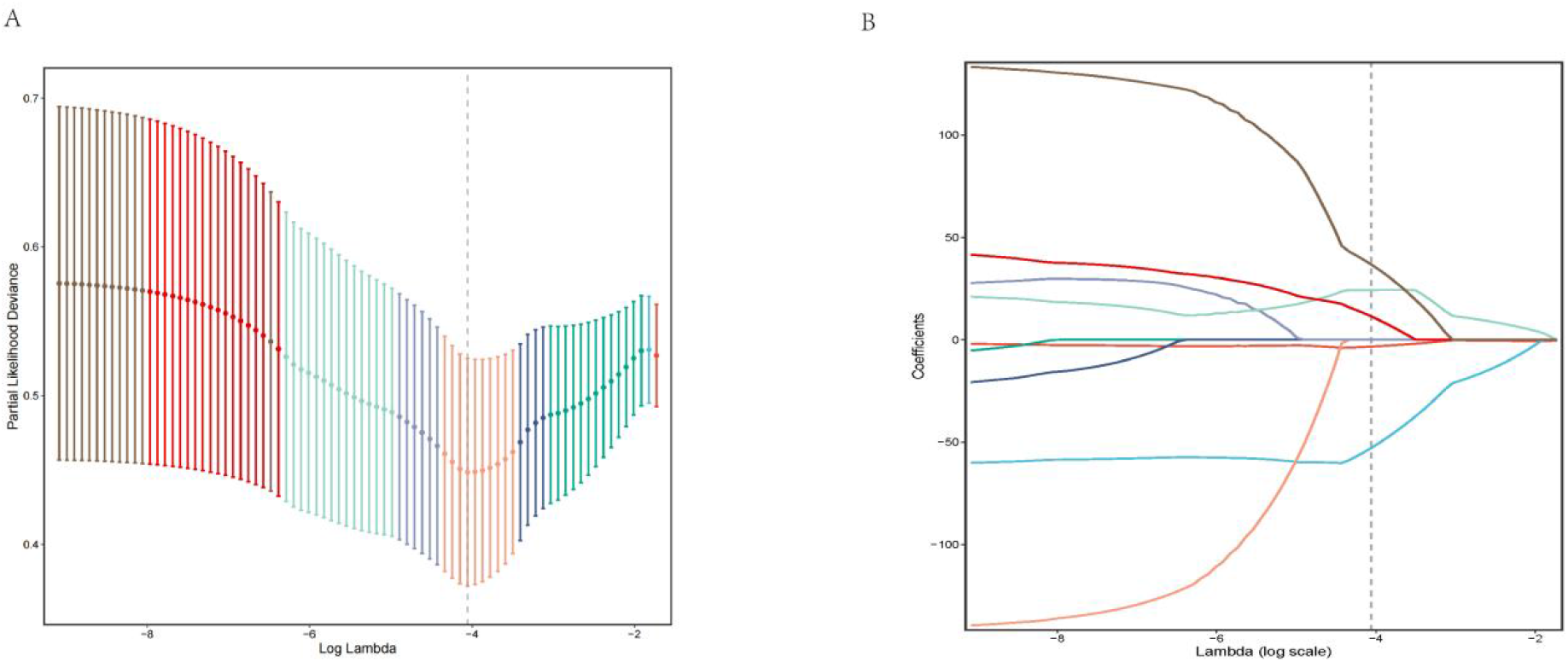
The LASSO regression analysis for potential predictors. (A)Selection of the tuning parameter (lambda) of the deviation in the LASSO regression based on the minimum criteria (dotted line). (B) A coefficient profile plot was generated against the log (lambda) sequence. LASSO, least absolute shrinkage and selection operator.

**Figure 3.**
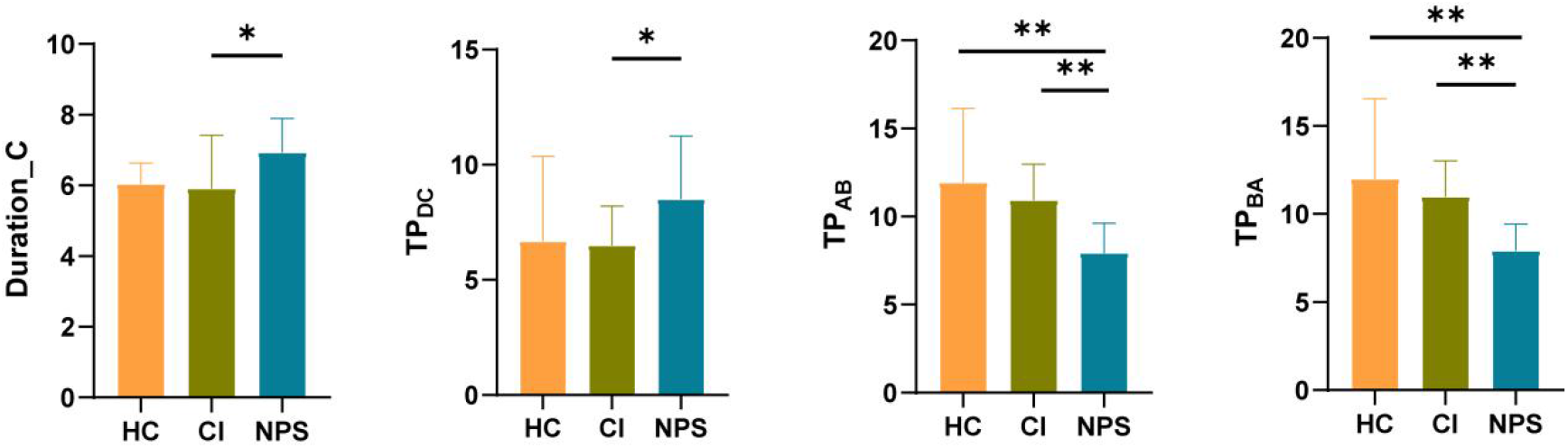
Potential predictors selected by the LASSO regression.

#### 3.3.2 Performance of the diagnostic model

Based on the LASSO results, a binary logistic model based on the 4 predictors for distinguishing NPS from CI was established. The model showed a high predictive value, with an AUC of 0.905 (95% CI: 0.784-1.000), a sensitivity of 100.0%, and a specificity of 76.9%. These results demonstrate that this model based on EEG microstate parameters had good diagnostic efficacy for identifying NPS from CI (**see Figure 4**).

**Figure 4.**
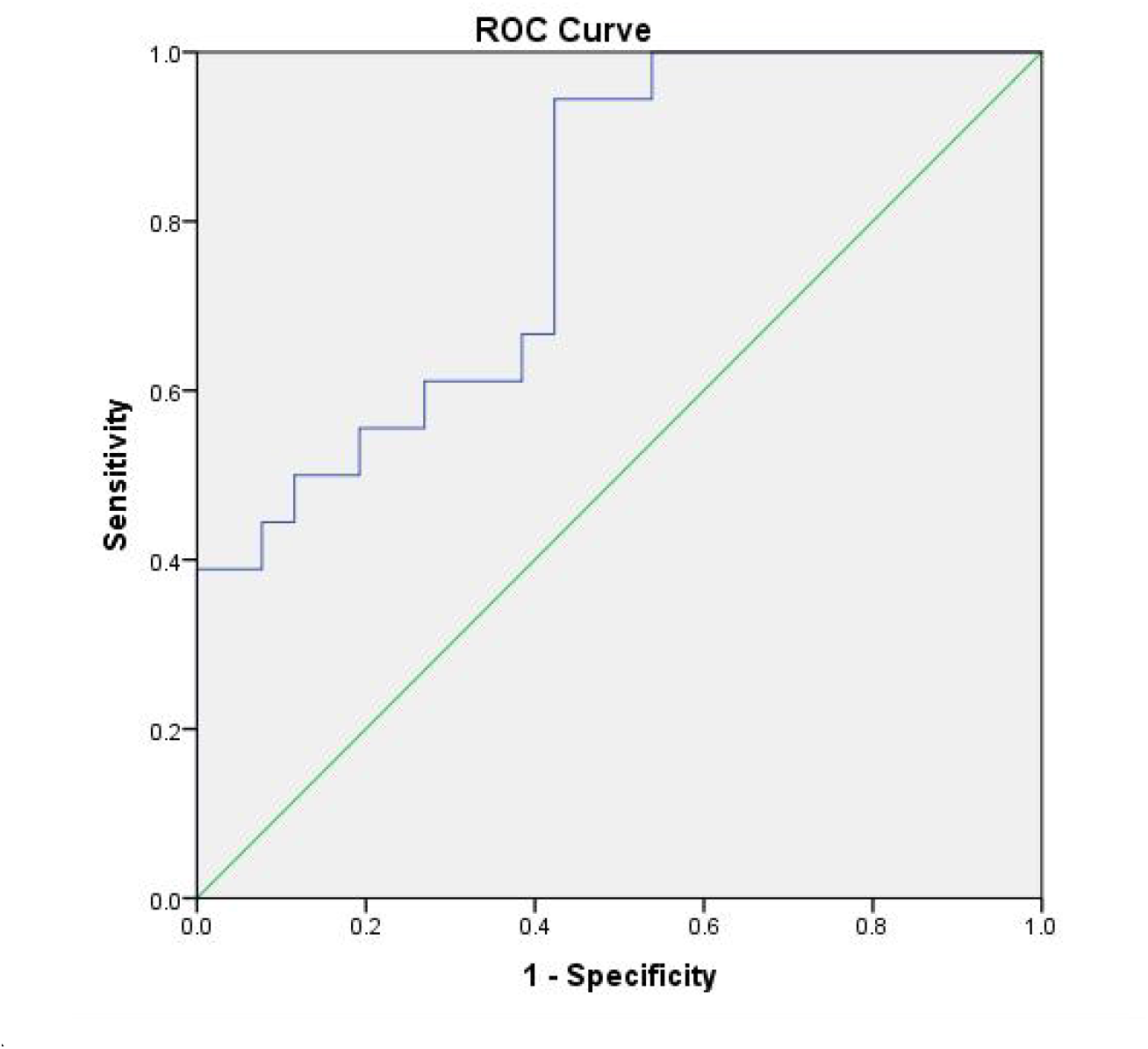
ROC curve for identifying NPS patients from CI patients.

## 4. Discussion

In this study, the basic information, neuropsychological assessment, MRI, and EEG microstate parameters among the three groups were compared. The result showed 8 differential EEG microstates between CI and NPS, including duration of class C, TP_BA_, TP_DC_, △TP_CD_, and △TP_DC,_ TP_AB_, △TP_AB_ and △TP_BA_, and 6 differential EEG microstates between CI and HC, including TP_BA_, △TP_CD_, △TP_DC_, △TP_AB_, △TP_AB_ and △TP_BA._ These results provide crucial insights into the abnormalities involved in neural networks alterations in patients with CI and NPS. Furthermore, a LASSO-logistic regression model for distinguishing NPS from CI patients was constructed, which included the duration of class C, TP_DC_, TP_AB_ and TP_BA_. The ROC results showed that this diagnostic model achieved AUC of 0.905 (95% CI: 0.784-1.000), with a sensitivity of 100.0% and a specificity of 76.9%. The diagnostic model based on EEG microstate parameters showed a good performance for differentiating NPS from CI patients.

EEG microstates are key indicators of transient states of global neuronal synchrony, and are characterized by stable scalp voltage topographies lasting only a few milliseconds. These EEG microstates signify orderly transitions and coordinated changes in large-scale neuronal activity, and are crucial for understanding brain function [29]. Previous studies have categorized EEG microstates into 4 classes, A, B, C, and D, each of which correlates with different functional networks. For instance, Class A is associated with inward thinking and memory processing and is likely tied to the default mode network (DMN), which is active during rest. Class B is related to sensory processing, and may be connected to the sensorimotor network. Class C is also linked to the DMN and self-reflection, whereas class D is correlated with emotional processing, possibly through the limbic network [30]. Thus, variations in different types of EEG microstate parameters may reflect abnormalities in specific resting-state functional networks.

In this study, compared to CI participants, the duration of class C and TP _DC_ are significantly longer in NPS patients. The increased TP_DC_ is believed to contribute to an increase in the duration of class C, which suggested that the increased duration of class C may be an important feature of NPS patients. The possible reason is that abnormal changes in class C are related to functional disruption of the DMN [29, 31]. The DMN is a distributed network that is active during task-negative/idle states, and consists of the medial prefrontal cortex, posterior cingulate cortex, hippocampal formation, and lateral temporal cortex [32]. Previous studies have shown that the DMN is not only involved in memory [33] but also plays a key role in social and affective behaviors [34]. Therefore, duration of class C may also be associated with affective symptoms resulting from DMN dysfunction. Furthermore, in the early stages of AD progression, Aβ accumulation first occurs within the DMN area, thereby affecting the functional connectivity of that area [35, 36], and the increased sedimentation of Aβ in the brain is also positively correlated with the severity of NPS [37-39]. Therefore, the abnormally increased duration of class C may reflect a significant increase in Aβ load in the brain and subsequent disruption of DMN function in patients with NPS. This finding suggests the possibility of a common mechanism between CI and NPS. However, to date, no study has directly demonstrated a relationship between the class C microstate and the structure and function of the DMN in NPS patients. Therefore, this conclusion requires further validation through synchronized fMRI-EEG studies.

The TP between microstate parameters not only provides information on changes in brain electrical activity patterns but also provides an important perspective for understanding how the brain dynamically regulates different cognitive and emotional states [40]. In this study, compared with HC, the TP between A and B decreased in CI patients, which showed a significant gradient like increase in NPS patients, indicating a possible overlap mechanism between CI and NPS phenotype of AD. This is consistent with most previous research findings that NPS and AD patients shared abnormalities in brain network function[41]. However, direct evidence of the presence of TPs between EEG microstates in AD patients with NPS has not yet well reported. Based on the original function of the microstate classes, our results could be explained by that the decreased TP between A and B may represent the sequence of networks that constitute large-scale brain networks. Disturbance in such a structure of network operations may result in disconnection between brain networks, which thereby leads to dysfunctional behavior.

## Limitations

There are several limitations in this study. First, the sample size was limited, and we only have single center data which containing NPS, CI and HC. However, we used LASSO regression to reduce the over-fitting and improve the stability of the results. This conclusion need to be replicate in further large-sample size study. Furthermore, this study is a cross-sectional study, only the diagnostic performance of EEG microstate parameters for distinguishing NPS from CI was analyzed, and their predictive performance for NPS and CI needs further validation in longitudinal studies. Additionally, 4 microstate classes were identified, which were considered the most robust classification, may affect performance of model. Therefore, future work should also evaluate whether it is possible to use more than 4 microstate classifications to construct the model.

## Conclusion

In summary, this study found that 8 differential EEG microstates between CI and NPS and 6 differential EEG microstates between CI and HC participants. These results provide crucial insights into the abnormalities involved in neural networks alterations in patients with CI and NPS. Based on EEG microstate, a LASSO-logistic model was constructed, which achieved high performance for distinguishing NPS from CI patients. This results could provide guidance for clinical assessment and use of drugs in AD patients. Overall speaking, this work suggests EEG microstates could be a potential imaging biomarker for NPS identification in AD patients.

## Data Availability

Availability of data and materials
The data that support the findings of this study are available from the corresponding author on reasonable request.

## Declarations

### Ethical Approval

Not applicable.

### Authors’ contributions

Lihong Huang designed the study; Zhangjing Deng,Mengyu Yan, and Lihong Huang collected the data; Lihong Huang analyzed the data; Lihong Huang organized the papers and wrote the manuscript; and Weihua Yu and Yang Lü reviewed and edited the manuscript. All the authors have read and approved the final manuscript.

### Availability of data and materials

The data that support the findings of this study are available from the corresponding author on reasonable request.

